# Aging measures and cancer: Findings from the Health and Retirement Study

**DOI:** 10.1101/2023.09.20.23295845

**Authors:** Shuo Wang, Anne Prizment, Puleng Moshele, Sithara Vivek, Anne H. Blaes, Heather H. Nelson, Bharat Thyagarajan

## Abstract

**Background:** Compared to cancer-free persons, cancer survivors of the same chronological age (CA) have increased physiological dysfunction, i.e., higher biological age (BA), which may lead to higher morbidity and mortality. We estimated BA using eight aging metrics: BA computed by Klemera Doubal method (KDM-BA), phenotypic age (PhenoAge), five epigenetic clocks (ECs, Horvath, Hannum, Levine, GrimAge, and pace of aging (POA)), and subjective age (SA). We tested if aging constructs were associated with total cancer prevalence and all-cause mortality in cancer survivors and controls, i.e., cancer-free persons, in the Health and Retirement Study (HRS), a large population-based study.

**Methods:** In 2016, data on BA-KDM, PhenoAge, and SA were available for 946 cancer survivors and 4,555 controls; data for the five ECs were available for 582 cancer survivors and 2,805 controls. Weighted logistic regression was used to estimate the association between each aging construct and cancer prevalence (odds ratio, OR, 95%CI). Weighted Cox proportional hazards regression was used to estimate the associations between each aging construct and cancer incidence as well as all-cause mortality (hazard ratio, HR, 95%CI). To study all BA metrics (except for POA) independent of CA, we estimated age acceleration as residuals of BA regressed on CA.

**Results:** Age acceleration for each aging construct and POA were higher in cancer survivors than controls. In a multivariable-adjusted model, five aging constructs (age acceleration for Hannum, Horvath, Levine, GrimAge, and SA) were associated with cancer prevalence. Among all cancer survivors, age acceleration for PhenoAge and four ECs (Hannum, Horvath, Levine, and GrimAge), was associated with higher all-cause mortality over 4 years of follow-up. PhenoAge, Hannum, and GrimAge were also associated with all-cause mortality in controls. The highest HR was observed for GrimAge acceleration in cancer survivors: 2.03 (95% CI, 1.58-2.60). In contrast, acceleration for KDM-BA and POA was significantly associated with mortality in controls but not in cancer survivors. When all eight aging constructs were included in the same model, two of them (Levine and GrimAge) were significantly associated with mortality among cancers survivors. None of the aging constructs were associated with cancer incidence.

**Conclusion:** Variations in the associations between aging constructs and mortality in cancer survivors and controls suggests that aging constructs may capture different aspects of aging and that cancer survivors may be experiencing age-related physiologic dysfunctions differently than controls. Future work should evaluate how these aging constructs predict mortality for specific cancer types.

## INTRODUCTION

One of the strongest risk factors for cancer occurrence is chronological age (CA), such that older individuals are at much higher risk for cancer than younger individuals.^1,2^ Compared to cancer-free individuals, cancer survivors have more physiological dysfunctions, which may be caused by cancer treatment, the body’s response to cancer (e.g., immunologic), the effects of cancer on the body (e.g., cachexia), the presence of unhealthy lifestyle factors, or an interaction between these factors.^3^ Understanding the aging process in cancer survivors may lead to improvements in outcomes through interventions that slow down physiological dysfunction (i.e., lifestyle changes or pharmacologic treatments).

Because individuals with the same CA may have very different physiologic and psychosocial dysfunctions, the term biological age (BA) has been introduced. BA estimates the extent of accumulation of physiological damage in individuals with the same CA. To estimate BA, multiple aging constructs have been developed including epigenetic clocks (ECs) based on DNA methylation profiles,^4-7^ proteomic aging clocks (using circulating proteins),^8,9^ transcriptomic clocks (using gene expression data),^10-12^ and multidomain aging constructs comprised of biochemical/hematological/physical markers. Consistent with the hypothesis that cancer survivors have increased physiological dysfunction, several recent studies have found that compared to similarly aged people without cancer, BA was higher among cancer survivors. For instance, two ECs developed by Horvath and Hannum were higher among cancer survivors in NHANES^13^ and an EC developed by Levine was higher among survivors of breast cancer in the Thinking and Living with Cancer (TLC) study.^14^ Higher EC has also been associated with an increased risk of mortality.^13,15^ In addition, different ECs adjusted for CA, were associated with incidence of cancer, including total,^16,17^ lung,^5,18^ breast,^19,20^ and pancreatic cancers,^21^ but the associations were inconsistent across aging metrics and different studies.

In addition to ECs, two multidomain measures-based aging constructs: a BA metrics based on biomarker and physiological measures and estimated by the Klemara Doubal algorithm (KDM-BA)^22-24^ and phenotypic age (PhenoAge) based on biomarkers associated with mortality^5,24-27^ have been developed, though their utility among cancer survivors has not been evaluated in previous studies. In UK Biobank, a large prospective cohort study, age-adjusted KDM-BA and PhenoAge have been associated with the risk of total, lung and colorectal cancers, while PhenoAge was additionally associated with breast cancer risk.^24^ In addition to biomarker-based aging measures, subjective age (SA), which estimates how old people feel, i.e., an individual’s self-perception of their own age,^28-30^ was 2% higher than CA in people diagnosed with cancer and 8% younger than CA among cancer-free controls.^31^ Moreover, higher SA was shown to be correlated with worse qualify of life in cancer survivors.^30,31^ In our prior work, we have shown that higher SA is associated with adverse biomarker profiles,^29^ and other studies have reported that SA is associated with negative health outcomes including higher mortality in the general population.^28,32,33^

Although measures of BA, such as PhenoAge, KDM-BA, ECs, and SA, are correlated with each other, they likely estimate different aspects of the underlying age-related physiological dysfunction. However, to date, there has been limited data to evaluate the relative utility of the various measures of BA in cancer survivors. Hence, we examined whether eight aging metrics including KDM-BA, PhenoAge, SA, and five ECs (Horvath, Hannum, Levine, GrimAge, and pace of aging (POA)) were higher in cancer survivors compared to cancer-free individuals in the Health and Retirement Study (HRS), a large population-based study. We also examined the associations of these measures with all-cause mortality among cancer survivors and cancer-free individuals. Finally, we conducted an exploratory analysis to evaluate whether any of these aging constructs were associated with total incident cancer risk in participants followed for four years.

## METHODS

### Study Population

The HRS study is an ongoing biennial longitudinal study of a nationally representative sample of individuals over 50 years old in the United States, starting from 1992. At each survey, participants had either a face-to-face interview or an interview over the phone to report their demographics, lifestyle, and medical history. SA data was obtained by responding to the question “How old do you feel?” for half of all participants in 2014 and for the other half in 2016. The biomarker data used to compute KDM-BA and PhenoAge were measured in blood samples collected from participants who completed a survey and blood collection in 2016 (N = 9,193). DNA methylation profiles were measured in a nonrandom subsample of participants (N = 4,018) who attended the HRS 2016 Venous Blood Study.^34^ A detailed description of the blood sample collection and processing is available on the HRS study website.^34^

### Biological age constructs

#### Biological Age estimated using Klemera-Doubal method (KDM-BA)

KDM-BA was calculated using 10 clinical markers that represent the decline in age-related physiological functioning and susceptibility to disease in old age (Supplemental Table 1).^22^ These 10 markers include systolic blood pressure (SBP), total cholesterol, fasting glucose, cytomegalovirus infection (CMV), C-Reactive Protein (CRP), serum creatinine, blood urea nitrogen (BUN), alkaline phosphatase, albumin, and peak flow measurement.^22^ The KDM-BA in this study was computed using R package “BioAge” and the in-built “kdm_calc” function.^35^

#### Phenotypic Age (PhenoAge)

PhenoAge was computed as a weighted sum of nine biomarkers (albumin, creatinine, glucose, log-transformed CRP (log CRP), lymphocyte percent, mean cell volume, red blood cell distribution width, alkaline phosphatase, and white blood cell count) as well as a person’s CA (all the variables were modeled as continuous)^5,25^ (Supplemental Table 1). PhenoAge in this study was computed using R package “BioAge” and the in-built “phenoage_calc” function.^35^

#### Epigenetic Clocks (ECs)

Thirteen published ECs were calculated in HRS.^36^ Among those 13 ECs, we selected five ECs that are commonly used in cancer studies: Horvath,^37^ Hannum,^4^ and Levine ECs,^5^ GrimAge^17^, and pace of aging (POA), which is measured using DunedinPoAm38^7^ (Supplemental Table 1). Horvath, Hannum, and Levine ECs, and GrimAge were expressed in years while POA was measured in “years of physiological decline occurring per 12 months of calendar time”^7^ (Supplemental Table 1).

### Ascertainment of cancer and mortality

Cancer survivors were defined as participants who answered “yes” to the question “Has a doctor ever told you that you have cancer or a malignant tumor, excluding minor skin cancer?” on the 2016 survey. Participants who answered “no” to the above question were considered as controls, i.e., participants without cancer. The self-reported cancer-related information collected in the 2016 survey was supplemented with information on site and diagnosis date that were reported in all surveys conducted from 1992 to 2016. Among controls in 2016, we used the 2018 and 2020 surveys to identify incident cancer cases up to 2020. Participants who reported a new cancer diagnosis in either the 2018 or 2020 surveys were further asked to report the month and year of their most recent cancer diagnosis. The earlier date reported at either survey was considered as the date of cancer diagnosis. For all participants, month and year of death was derived from interviews provided by household members.

### Characteristics of interest

The following characteristics were obtained from the 2016 survey: CA (in years), sex (female/male), race/ethnicity (Non-Hispanic White, Non-Hispanic Black, Hispanic White, Hispanic Black, or Other), and smoking status (self-reported; current, former, or never smokers). A comorbidity index was constructed using seven self-reported conditions diagnosed by a physician. These conditions included hypertension, lung disease, cardiac disorders, stroke, arthritis, diabetes, and psychiatric problems. The measure of CMV seroprevalence was described previously^34^ and was reported as nonreactive [<0.5 COI (cutoff interval)), borderline (0.5 to <1.0 COI), or reactive (≥1.0 COI]. In this study, CMV infection was used as a binary variable, with the borderline and nonreactive CMV groups combined. Height was reported in inches and weight was reported in pounds. Height and weight data were obtained for half of all participants in 2014 and for the other half in 2016. BMI was calculated as Weight in lbs/ (Height in inches)^2^ *703.

### Statistical Analysis

The analyses described below were conducted using SAS version 9.4. All the analyses were adjusted for survey weights to yield nationally representative estimates.

The analyses were conducted in two samples: Sample A included participants with the data on SA, KDM-BA, and PhenoAge, and Sample B included participants with data on ECs. Sample A excluded participants with missing values for any clinical biomarker that comprised KDM-BA or PhenoAge, as well as those who did not answer questions about SA and those without survey weights, characteristics of interest, cancer status, or vital status. This resulted in 5,501 participants, which included 946 cancer survivors and 4,555 controls, in Sample A. Sample B excluded participants without data on ECs, and those without survey weights, characteristics of interest, cancer status, or vital status. This resulted in 3,387 participants, which included 582 cancer survivors and 2,805 controls, in Sample B.

Because these aging constructs, except for POA, were correlated with CA (Supplemental Tables 2 and 3), we estimated age acceleration (abbreviated as Accel) for each of aging construct as residuals after regressing aging construct on CA to evaluate the effects of aging construct independent of age. The distributions of aging constructs and characteristics of interest were examined as weighted mean (standard deviation (SD)) or weighted percent across cancer status in 2016.

We used weighted logistic regression to estimate weighted odds ratios (ORs) and 95% confidence intervals (CIs) for the associations between each aging construct and cancer prevalence (cancer survivors vs. controls). We applied weighted Cox proportional hazard regression to examine association between each aging construct and all-cause mortality among cancer survivors and among controls. The follow-up for mortality started from the date of blood collection in 2016 and ended on either death or December 31, 2020, whichever occurred first.

The proportional hazard assumption, examined by including an interaction term between follow-up time and each of the aging construct was not violated in any of the models. In all main analyses, we constructed two models. Model 1 was adjusted for age and Model 2 was additionally adjusted for sex, race/ethnicity, BMI, smoking status, comorbidity index, and CMV infection. To ensure that active cancer treatment or undiagnosed cancer at the time of blood draw was not a major contributor to our findings, we conducted a sensitivity analysis that excluded participants whose cancer diagnosis was within two years of the blood collection.

We also performed three exploratory analyses. In the first exploratory analysis, we investigated whether the association with mortality in cancer survivors was modified by sex or time since cancer diagnosis. We examined the associations with mortality among cancer survivors, stratified by sex and time since cancer diagnosis (<2 years, 2 to 5 years, and >5 years). We did not examine associations stratified by race/ethnicity due to the limited number of deaths in Hispanic White and Non-Hispanic and Hispanic Black participants. In the second exploratory analysis, we investigated the associations with total cancer incidence among controls, i.e., cancer-free participants in 2016, by applying weighted Cox proportional hazard regression. The follow-up for incident cancer started from the date of blood collection in 2016 and ended on the date of cancer diagnosis, death, loss to follow up, or December 31, 2020, whichever occurred first. Finally among participants who were in both Sample A and Sample B, we explored the correlations between each aging construct and CA, and between the age acceleration for each construct and CA. Subsequently, we included all aging constructs in a single model simultaneously and evaluated their associations with mortality in cancer survivors and controls who were in both Sample A and Sample B.

## RESULTS

### Distributions of participant characteristics

Table 1 shows the distributions of participants’ characteristics by cancer status. Compared to controls, cancer survivors were more likely to be chronologically older. Cancer survivors also tended to be Non-Hispanic White and former smokers and to have a higher comorbidity index (Table 1). The correlations between aging constructs and CA were 0.03-0.92 while the age acceleration for each aging construct was not correlated with CA (Supplemental Tables 2 and 3).

**Table 1.**
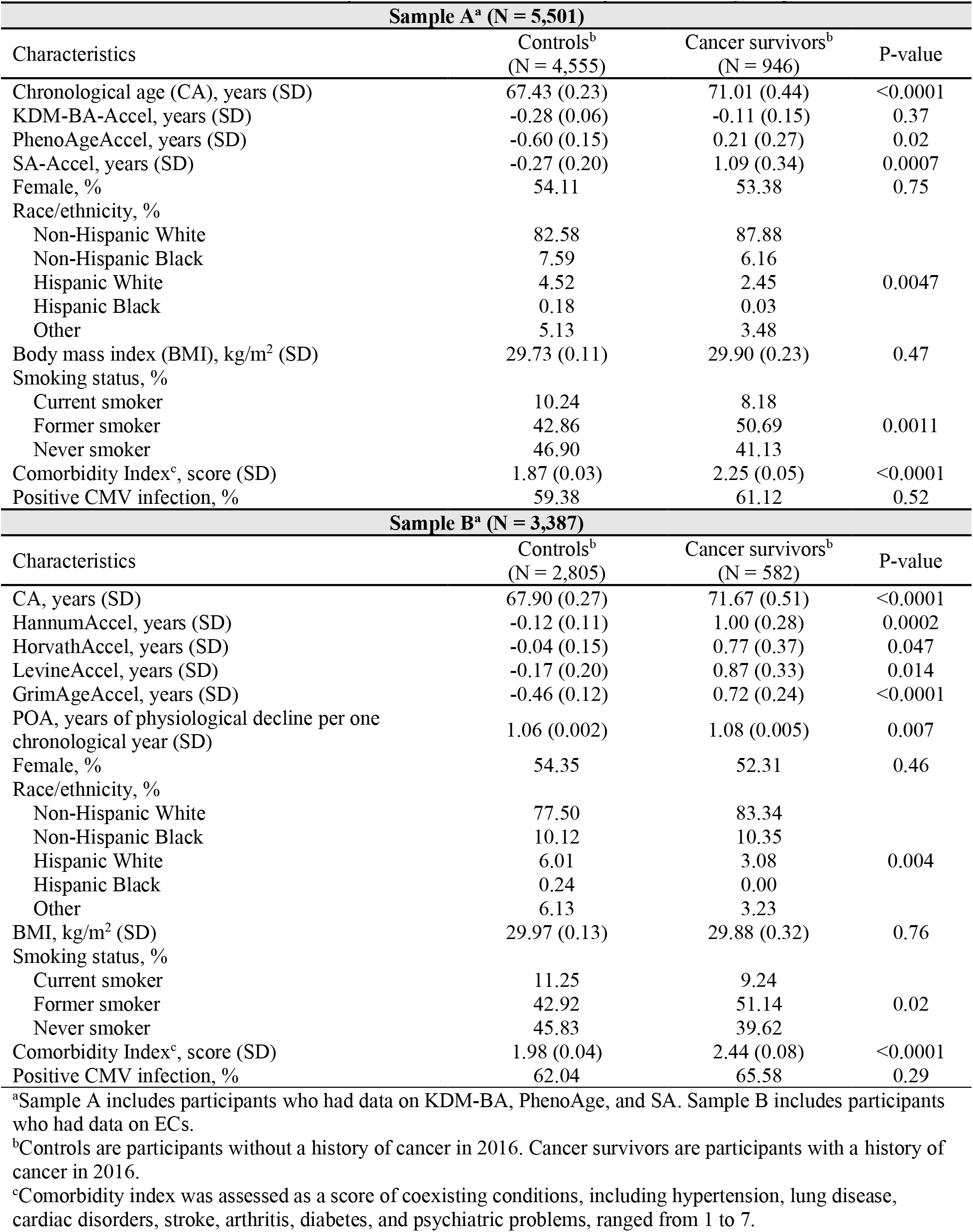
Distribution of characteristics by cancer status in 2016 after adjusted for survey weights, HRS.

### Associations between aging constructs and cancer prevalence

Age acceleration for each aging construct and POA was higher in cancer survivors than in controls (Table 1). In the fully adjusted model (Model 2), age acceleration for SA [OR (95% CI) per 1 SD=1.12 (1.03-1.21)], Hannum [1.24 (1.12-1.38)], Levine [1.14 (1.01-1.28)], and GrimAge [1.31 (1.13-1.52)] was significantly associated with cancer prevalence, but not for KDM-BA, PhenoAge, Horvath or POA (Table 2).

**Table 2.**
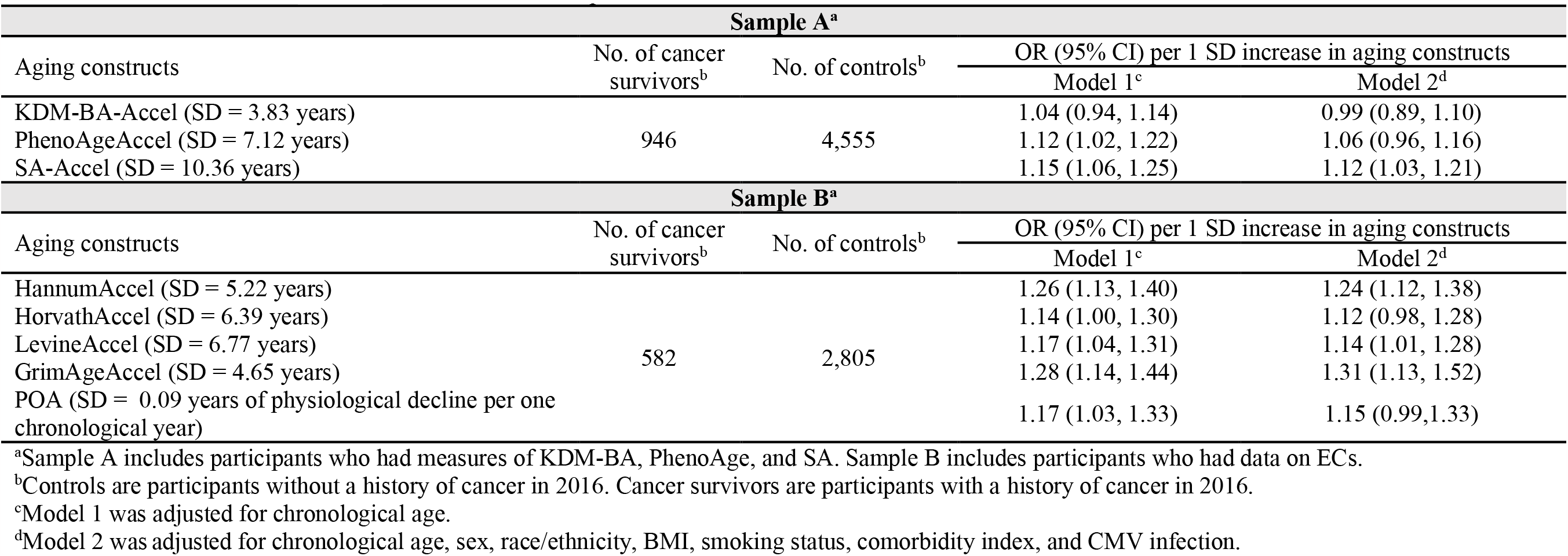
Associations between aging constructs and cancer prevalence in 2016, HRS.

### Associations between aging constructs and all-cause mortality in cancer survivors and controls

By 2020, 122 cancer survivors and 249 controls died in Sample A and 103 cancer survivors and 213 controls died in Sample B. In the fully adjusted model, PhenoAgeAccel, HannumAccel, and GrimAgeAccel were associated with mortality in both cancer survivors and controls with a higher HR estimate observed in cancer survivors (Table 3). For example, for GrimAge: hazard ratio (HR) (95% CI) per 1 SD=2.13 (1.52-2.99) in cancer survivors and 1.45 (1.19-1.78) in controls (Table 3). HorvathAccel [1.26 (1.02-1.55)] and LevineAccel [1.58 (1.29-1.94)] were significantly associated with mortality among cancer survivors only, while KDM-BA-Accel [1.32 (1.10-1.59)] and POA [1.31 (1.14-1.52)] were significantly associated with morality among controls only (Table 3). SA-Accel was not associated with mortality in either cancer survivors or controls (Table 3).

**Table 3.** Associations between aging constructs and mortality among cancer survivors and controls after adjusted for survey weights, HRS (2016-2020)

| Sample A<sup>a</sup> |  |  |  |  |
| --- | --- | --- | --- | --- |
| Cancer survivors (N = 946)<sup>b</sup> |  |  |  |  |
| Aging constructs | No. of deaths | Total person-year | HR (95% CI)<sup>c</sup> per 1 SD increase in aging constructs |  |
|  |  |  | Model 1<sup>c</sup> | Model 2<sup>d</sup> |
| KDM-BA-Accel (SD = 4.07 years) | 122 | 3,775 | 1.18 (0.99, 1.39) | 1.14 (0.95, 1.37) |
| PhenoAgeAccel (SD = 7.22 years) |  |  | 1.47 (1.17, 1.85) | 1.47 (1.14, 1.90) |
| SA-Accel (SD = 10.29 years) |  |  | 0.98 (0.68, 1.41) | 0.93 (0.64, 1.33) |
| Controls (N = 4,555)<sup>b</sup> |  |  |  |  |
| Aging constructs | No. of deaths | Total person-year | HR (95% CI)<sup>c</sup> per 1 SD increase in aging constructs |  |
|  |  |  | Model 1<sup>c</sup> | Model 2<sup>d</sup> |
| KDM-BA-Accel (SD = 3.78 years) | 249 | 18,861 | 1.39 (1.20, 1.60) | 1.32 (1.10, 1.59) |
| PhenoAgeAccel (SD = 7.09 years) |  |  | 1.43 (1.27, 1.61) | 1.37 (1.19, 1.58) |
| SA-Accel (SD = 10.37 years) |  |  | 1.22 (0.96, 1.54) | 1.18 (0.96, 1.45) |
| Sample B<sup>a</sup> |  |  |  |  |
| Cancer survivors (N = 582)<sup>b</sup> |  |  |  |  |
| Aging constructs | No. of deaths | Total person-year | HR (95% CI)<sup>c</sup> per 1 SD increase in aging constructs |  |
|  |  |  | Model 1<sup>c</sup> | Model 2<sup>d</sup> |
| HannumAccel (SD = 5.48 years) | 103 | 2,270 | 1.24 (1.07, 1.43) | 1.33 (1.15, 1.53) |
| HorvathAccel (SD = 7.01 years) |  |  | 1.19 (0.98, 1.49) | 1.26 (1.02, 1.55) |
| LevineAccel (SD = 6.88 years) |  |  | 1.49 (1.25, 1.77) | 1.58 (1.29, 1.94) |
| GrimAgeAccel (SD = 4.68 years) |  |  | 2.03 (1.58, 2.60) | 2.13 (1.52, 2.99) |
| POA (SD = 0.09 years of physiological decline per one chronological year) |  |  | 1.38 (1.12, 1.71) | 1.27 (0.97, 1.67) |
| Controls (N = 2,805)<sup>b</sup> |  |  |  |  |
| Aging constructs | No. of deaths | Total person-year | HR (95% CI)<sup>c</sup> per 1 SD increase in aging constructs |  |
|  |  |  | Model 1<sup>c</sup> | Model 2<sup>d</sup> |
| HannumAccel (SD = 5.14 years) | 213 | 11,439 | 1.20 (1.03, 1.40) | 1.19 (1.01, 1.41) |
| HorvathAccel (SD = 6.25 years) |  |  | 1.07 (0.88, 1.31) | 1.05 (0.85, 1.30) |
| LevineAccel (SD = 6.74 years) |  |  | 1.14 (0.95, 1.37) | 1.09 (0.90, 1.33) |
| GrimAgeAccel (SD = 4.63 years) |  |  | 1.57 (1.37, 1.82) | 1.45 (1.19, 1.78) |
| POA (SD = 0.09 years of physiological decline per one chronological year) |  |  | 1.45 (1.27, 1.67) | 1.31 (1.14, 1.52) |

<sup>a</sup>Sample A includes participants who had measures of KDM-BA, PhenoAge, and SA. Sample B includes participants who had data on ECs.
<sup>b</sup>Controls are participants without a history of cancer in 2016. Cancer survivors are participants with a history of cancer in 2016.
<sup>c</sup>Model 1 was adjusted for chronological age.
<sup>d</sup>Model 2 was adjusted for chronological age, sex, race/ethnicity, BMI, smoking status, comorbidity index, and CMV infection.

In the sensitivity analysis, after excluding participants who developed cancer within two years of blood collection, the results for the associations with cancer prevalence and mortality were comparable to the results in our main analyses (Supplemental Tables 4 and 5).

In the exploratory analysis of interaction, gender statistically modified the association between GrimAgeAccel and mortality in cancer survivors (p-interaction = 0.002), with a stronger association observed in females (Supplemental Table 6). Time since cancer diagnosis did not modify the associations between any of the aging constructs and mortality in cancer survivors (Supplemental Table 7). In the exploratory analysis of cancer incidence, cancer diagnoses were self-reported by 182 participants in Sample A and by 122 participants in Sample B. None of the eight aging constructs were associated with cancer risk (Supplemental Table 8). In the last exploratory analysis, among the participants who were in both Sample A and Sample B, the correlations between eight aging constructs were 0.06-0.84 and the correlations between age acceleration for the aging constructs were 0.004-0.63 (Supplemental Table 9). When including all eight aging constructs in the same model, LevineAccel and GrimAgeAccel were significantly associated with mortality among cancer survivors and KDM-BA-Accel was significantly associated with mortality among controls (Supplemental Table 10).

## DISCUSSION

This study examined the association of BA, a measure of accumulated life course changes in biological systems, with cancer prevalence and all-cause mortality in a large nationally representative sample. A recent review^38^ summarized the common biomarkers used to estimate BA in cancer survivors such as P16^Ink4a^, telomere length, ECs, or allostatic load. In our study, we tested eight well-validated aging constructs that have been used in previous studies: BA estimated by the Klemara Doubal algorithm (KDM-BA), phenotypic age (PhenoAge), SA, as well as five ECs including Horvath, Hannum, Levine, GrimAge, and POA.

Our study found that cancer survivors had higher age acceleration and POA than controls and age acceleration for four metrics (SA, Hannum EC, Levine EC, and GrimAge) was significantly associated with cancer prevalence in the fully adjusted model. Among cancer survivors, age acceleration for PhenoAge and four ECs (Hannum, Horvath, Levine, and GrimAge) was associated with higher all-cause mortality over 4 years of follow-up. Cancer survivors may experience accelerated aging due to both cancer itself and cytotoxic cancer treatment, particularly radiation therapy and chemotherapy, which may cause epigenetic changes and cellular senescence features such as telomere shortening and alterations in DNA repair genes.^39,40^

In turn, these changes individually or in combination contribute to accelerated aging phenotypes in cancer survivors such as PhenoAge, KDM-BA or ECs. Consistent with our results, the Melbourne Collaborative Cohort study showed that per 5-year increment in age acceleration for Horvath and Hannum ECs was significantly associated with a 4-6% increased risk of all-cause death in cancer survivors.^15^ Further, a recent NHANES study reported an increased risk of all-cause death associated with the upper quartile of the Levine EC (HR=3.04, 95%CI: 2.31-4.00).^13^ The physiological changes among cancer survivors along with stress of managing cancer, worries about cancer recurrence, life changes, job and family issues resulting from having cancer may contribute to a greater SA in cancer survivors. Consistent with the mechanism, our study found cancer survivors on average had age acceleration for SA of 1.09 years, while controls on average had age acceleration for SA of -0.27 years. In our study, SA, which is a non-biomarker-based measure, was correlated with CA (r=0.61) and with other biomarker-based aging constructs (r=0.06-0.52), but these correlations were weaker than for the biomarker-based constructs. We also found that age acceleration for SA was associated with cancer prevalence in a fully adjusted model, however, there was no association with mortally in cancer survivors. To our knowledge, there were no studies examining association between SA and mortality in cancer survivors. It is likely that SA captures different aspects of aging than biomarker-based aging constructs and may not accurately predict mortality in cancer survivors because they have a milieu of different age- and cancer-related issues that may not be captured by a one-time question about how one feels.

In our study, among controls, age acceleration for four aging constructs (KDM-BA, PhenoAge, Hannum EC, and GrimAge) as well as POA was associated with greater all-cause mortality. In agreement with our findings, higher KDM-BA and PhenoAge were associated with worse mortality in the NHANES and UK Biobank studies.^24,26,41^ We also found an increased HR for mortality associated with SA but not statistically significant. Increased HR in controls in our study is in line with several previous studies that reported associations between older SA and mortality in the general population.^32,33,42,43^

In our study, age acceleration for three constructs, PhenoAge, Hannum EC, and GrimAge, was associated with mortality in both cancer survivors and controls. The lack of associations with KDM-BA and POA in cancer survivors may be explained by a smaller sample size in cancer survivors than controls. However, we also found associations with mortality for the Horvath and Levine ECs in cancer survivors but not in controls, and the associations for the Hannum EC and GrimAge were stronger in cancer survivors. Stronger associations between aging constructs and mortality in cancer survivors support the conclusion from the recent NHANES study of PhenoAge in cancer survivors and controls in relation to all-cause mortality.^13^ Our findings suggest that different aging constructs may predict mortality differently in cancer survivors and in controls.

To compare relative performance of aging constructs, we included all eight aging constructs in a single model simultaneously. We found that Levine EC and GrimAge were independently associated with mortality among cancer survivors and KDM-BA was independently associated with mortality among controls. These results along with the low correlations between age acceleration of aging constructs (r=0.004-0.63) suggest that different aging constructs may capture different aspects of BA.

Aging and cancer may have a bidirectional association. Not only do cancer survivors have greater BA but persons with higher BA may be at increased cancer risk most likely because both cancer and aging result from genetic and epigenetic damage accumulation,^44^ inflammation, oxidative stress, and other damage.^45-47^ Contrary to the consistent associations of various aging constructs with prevalent cancer and mortality, there was no association with cancer incidence for any aging construct in our study. These findings do not agree with several previous studies that found associations of different ECs, such as Hannum, Horvath, GrimAge and Levine, with the incidence of total cancer,^17,48^ breast,^20,49-51^ lung,^18^ male colon cancer^49^ (summarized in a recent review^52^). Positive associations have been also reported of KDM-BA and PhenoAge with the incidence of total, lung and colorectal cancer in UK Biobank, a large prospective study.^24^ In that study, PhenoAge was also associated with breast cancer risk.^24^ Most likely, in our study, there was no association with the cancer risk due to a limited power and the inability to evaluate individual cancer types.

Limitations in our study include a self-reported information about cancer diagnosis and other cancer characteristics. However, a recent validation study found that, compared to Medicare data, self-report first incident cancer diagnoses in HRS had 73.2% sensitivity and 96.29% specificity.^53^ In addition, we have a limited sample size to examine associations in cancer survivors especially for ECs that have been measured in a smaller sample. The strengths included the population-based longitudinal study, the detailed information about various risk factors and comorbidities, and the assessment of different aging measures in the same study in both cancer survivors and controls.

Estimating BA in cancer survivors is particularly important because it may serve as a marker of survivor’s ability to manage the rigors of cancer treatment and their physiological function. BA may be a better marker than CA that is commonly used for cancer therapy guidelines because cancer survivors of the same CA have very different ability to tolerate treatment. Our study also suggests that, compared to a single aging construct, including multiple aging constructs may more comprehensively capture the global age-related dysfunction seen among cancer survivors and should be examined in the future studies.

## Supporting information

Supplemental Tabel 7

## Data Availability

The data used in the current study are available at: https://hrsdata.isr.umich.edu/data-products/public-survey-data and https://hrsdata.isr.umich.edu/data-products/sensitive-health.

## ACKNOWLEDGEMENT

This work was supported by R01AG060110 and R01CA267977. The Health and Retirement Study is supported by U01AG009740.

## Notes

### Competing Interest Statement

The authors have declared no competing interest.

## References

1. Guida JL, Ahles TA, Belsky D, et al. Measuring aging and identifying aging phenotypes in cancer survivors. JNCI: Journal of the National Cancer Institute. 2019;111(12):1245–1254.

2. Sierra F, Kohanski R. Geroscience and the trans-NIH GeroScience Interest Group. GSIG Geroscience 39 (1): 1–5. 2017.

3. Pluijm SMF. Accelerated Aging as a Paradigm to Understand the Late Effects of Cancer Therapies. Endocrine and Metabolic Late Effects in Cancer Survivors. 2021;54:1–9.

4. Hannum G, Guinney J, Zhao L, et al. Genome-wide methylation profiles reveal quantitative views of human aging rates. Mol Cell. Jan 24 2013;49(2):359–367. doi:10.1016/j.molcel.2012.10.016

5. Levine ME, Lu AT, Quach A, et al. An epigenetic biomarker of aging for lifespan and healthspan. Aging. 2018;10(4):573.

6. Horvath S. DNA methylation age of human tissues and cell types. Genome biology. 2013;14(10):3156.

7. Belsky DW, Caspi A, Arseneault L, et al. Quantification of the pace of biological aging in humans through a blood test, the DunedinPoAm DNA methylation algorithm. eLife. 2020;9:e54870.

8. Tanaka T, Basisty N, Fantoni G, et al. Plasma proteomic biomarker signature of age predicts health and life span. eLife. 2020;9:e61073.

9. Lehallier B, Shokhirev MN, Wyss-Coray T, Johnson AA. Data mining of human plasma proteins generates a multitude of highly predictive aging clocks that reflect different aspects of aging. Aging cell. 2020;19(11):e13256.

10. Meyer DH, Schumacher B. BiT age: A transcriptome-based aging clock near the theoretical limit of accuracy. Aging Cell. Mar 2021;20(3):e13320. doi:10.1111/acel.13320

11. Peters MJ, Joehanes R, Pilling LC, et al. The transcriptional landscape of age in human peripheral blood. Nat Commun. Oct 22 2015;6:8570. doi:10.1038/ncomms9570

12. Nakamura S, Kawai K, Takeshita Y, et al. Identification of blood biomarkers of aging by transcript profiling of whole blood. Biochem Biophys Res Commun. Feb 10 2012;418(2):313–8. doi:10.1016/j.bbrc.2012.01.018

13. Zhang D, Leeuwenburgh C, Zhou D, et al. Analysis of Biological Aging and Risks of All-Cause and Cardiovascular Disease-Specific Death in Cancer Survivors. JAMA Netw Open. Jun 1 2022;5(6):e2218183. doi:10.1001/jamanetworkopen.2022.18183

14. Rentscher KE, Bethea TN, Zhai W, et al. Epigenetic aging in older breast cancer survivors and noncancer controls: preliminary findings from the Thinking and Living with Cancer Study. Cancer. Sep 1 2023;129(17):2741–2753. doi:10.1002/cncr.34818

15. Dugué PA, Bassett JK, Joo JE, et al. DNA methylation-based biological aging and cancer risk and survival: Pooled analysis of seven prospective studies. Int J Cancer. Apr 15 2018;142(8):1611–1619. doi:10.1002/ijc.31189

16. Zheng Y, Joyce BT, Colicino E, et al. Blood Epigenetic Age may Predict Cancer Incidence and Mortality. EBioMedicine. Mar 2016;5:68–73. doi:10.1016/j.ebiom.2016.02.008

17. Lu AT, Quach A, Wilson JG, et al. DNA methylation GrimAge strongly predicts lifespan and healthspan. Aging (Albany NY). 2019;11(2):303.

18. Levine ME, Hosgood HD, Chen B, Absher D, Assimes T, Horvath S. DNA methylation age of blood predicts future onset of lung cancer in the women’s health initiative. Aging. 2015;7(9):690.

19. Sehl ME, Carroll JE, Horvath S, Bower JE. The acute effects of adjuvant radiation and chemotherapy on peripheral blood epigenetic age in early stage breast cancer patients. NPJ breast cancer. 2020;6(1):1–5.

20. Kresovich JK, Xu Z, O’Brien KM, Weinberg CR, Sandler DP, Taylor JA. Methylation-based biological age and breast cancer risk. JNCI: Journal of the National Cancer Institute. 2019;111(10):1051–1058.

21. Chung M, Ruan M, Zhao N, et al. DNA methylation aging clocks and pancreatic cancer risk: Pooled analysis of three prospective nested case-control studies. medRxiv. 2020;

22. Levine ME. Modeling the rate of senescence: can estimated biological age predict mortality more accurately than chronological age? Journals of Gerontology Series A: Biomedical Sciences and Medical Sciences. 2013;68(6):667–674.

23. Kuo CL, Pilling LC, Liu Z, Atkins JL, Levine ME. Genetic associations for two biological age measures point to distinct aging phenotypes. Aging Cell. Jun 2021;20(6):e13376. doi:10.1111/acel.13376

24. Mak JKL, McMurran CE, Kuja-Halkola R, et al. Clinical biomarker-based biological aging and risk of cancer in the UK Biobank. Br J Cancer. Jul 2023;129(1):94–103. doi:10.1038/s41416-023-02288-w

25. Liu Z, Kuo PL, Horvath S, Crimmins E, Ferrucci L, Levine M. A new aging measure captures morbidity and mortality risk across diverse subpopulations from NHANES IV: A cohort study. PLoS Med.Dec 2018;15(12):e1002718. doi:10.1371/journal.pmed.1002718

26. Liu Z, Chen X, Gill TM, Ma C, Crimmins EM, Levine ME. Associations of genetics, behaviors, and life course circumstances with a novel aging and healthspan measure: Evidence from the Health and Retirement Study. PLoS Med. Jun 2019;16(6):e1002827. doi:10.1371/journal.pmed.1002827

27. Thomas A, Belsky DW, Gu Y. Healthy lifestyle behaviors and biological aging in the US National Health and Nutrition Examination Surveys 1999-2018. The journals of gerontology Series A, Biological sciences and medical sciences. Mar 10 2023;doi:10.1093/gerona/glad082

28. Shinan-Altman S, Werner P. Subjective Age and Its Correlates Among Middle-Aged and Older Adults. Int J Aging Hum Dev. Jan 2019;88(1):3–21. doi:10.1177/0091415017752941

29. Thyagarajan B, Shippee N, Parsons H, et al. How Does Subjective Age Get “Under the Skin”? The Association Between Biomarkers and Feeling Older or Younger Than One’s Age: The Health and Retirement Study. Innovation in aging. Aug 2019;3(4):igz035. doi:10.1093/geroni/igz035

30. Boehmer S. Does felt age reflect health-related quality of life in cancer patients? Psychooncology. Aug 2006;15(8):726–38. doi:10.1002/pon.1011

31. Laryionava K, Schönstein A, Heußner P, Hiddemann W, Winkler EC, Wahl HW. Experience of Time and Subjective Age When Facing a Limited Lifetime: The Case of Older Adults with Advanced Cancer. J Aging Health. Aug-Sep 2022;34(4-5):736–749. doi:10.1177/08982643211063162

32. Stephan Y, Sutin AR, Terracciano A. Subjective Age and Mortality in Three Longitudinal Samples. Psychosom Med. Sep 2018;80(7):659–664. doi:10.1097/psy.0000000000000613

33. Kotter-Grühn D, Kleinspehn-Ammerlahn A, Gerstorf D, Smith J. Self-perceptions of aging predict mortality and change with approaching death: 16-year longitudinal results from the Berlin Aging Study. Psychol Aging. Sep 2009;24(3):654–67. doi:10.1037/a0016510

34. Crimmins E, Faul J, Thyagarajan B, Weir D. Venous blood collection and assay protocol in the 2016 Health and Retirement Study. Ann Arbor, MI: Survey Research Center, Institute for Social Research, University of Michigan. 2017;

35. Kwon D, Belsky DW. A toolkit for quantification of biological age from blood chemistry and organ function test data: BioAge. Geroscience. Dec 2021;43(6):2795–2808. doi:10.1007/s11357-021-00480-5

36. Crimmins EM, Thyagarajan B, Levine ME, Weir DR, Faul J. Associations of Age, Sex, Race/Ethnicity and Education with 13 Epigenetic Clocks in a Nationally Representative US Sample: The Health and Retirement Study. The Journals of Gerontology: Series A. 2021;

37. Horvath S. DNA methylation age of human tissues and cell types. Genome Biol. 2013;14(10):R115. doi:10.1186/gb-2013-14-10-r115

38. Mandelblatt JS, Ahles TA, Lippman ME, et al. Applying a Life Course Biological Age Framework to Improving the Care of Individuals With Adult Cancers: Review and Research Recommendations. JAMA Oncol. Nov 1 2021;7(11):1692–1699. doi:10.1001/jamaoncol.2021.1160

39. Wang S, Prizment A, Thyagarajan B, Blaes A. Cancer Treatment-Induced Accelerated Aging in Cancer Survivors: Biology and Assessment. Cancers (Basel). Jan 23 2021;13(3)doi:10.3390/cancers13030427

40. Guida JL, Agurs-Collins T, Ahles TA, et al. Strategies to Prevent or Remediate Cancer and Treatment-Related Aging. J Natl Cancer Inst. Apr 2020;doi:10.1093/jnci/djaa060

41. Parker DC, Bartlett BN, Cohen HJ, et al. Association of Blood Chemistry Quantifications of Biological Aging With Disability and Mortality in Older Adults. The journals of gerontology Series A, Biological sciences and medical sciences. Sep 16 2020;75(9):1671–1679. doi:10.1093/gerona/glz219

42. Rippon I, Steptoe A. Feeling old vs being old: associations between self-perceived age and mortality. JAMA Intern Med. Feb 2015;175(2):307–9. doi:10.1001/jamainternmed.2014.6580

43. Westerhof GJ, Miche M, Brothers AF, et al. The influence of subjective aging on health and longevity: a meta-analysis of longitudinal data. Psychol Aging. Dec 2014;29(4):793–802. doi:10.1037/a0038016

44. Campisi J. Aging, cellular senescence, and cancer. Annual review of physiology. 2013;75:685–705.

45. Il’yasova D, Colbert LH, Harris TB, et al. Circulating levels of inflammatory markers and cancer risk in the health aging and body composition cohort. Cancer Epidemiology and Prevention Biomarkers. 2005;14(10):2413–2418.

46. Zuo L, Prather ER, Stetskiv M, et al. Inflammaging and oxidative stress in human diseases: From molecular mechanisms to novel treatments. International journal of molecular sciences. 2019;20(18):4472.

47. Greten FR, Grivennikov SI. Inflammation and cancer: triggers, mechanisms, and consequences. Immunity. 2019;51(1):27–41.

48. Zheng Y, Joyce BT, Colicino E, et al. Blood epigenetic age may predict cancer incidence and mortality. EBioMedicine. 2016;5:68–73.

49. Durso DF, Bacalini MG, Sala C, et al. Acceleration of leukocytes’ epigenetic age as an early tumor and sex-specific marker of breast and colorectal cancer. Oncotarget. Apr 4 2017;8(14):23237–23245. doi:10.18632/oncotarget.15573

50. Ambatipudi S, Horvath S, Perrier F, et al. DNA methylome analysis identifies accelerated epigenetic ageing associated with postmenopausal breast cancer susceptibility. Eur J Cancer. Apr 2017;75:299–307. doi:10.1016/j.ejca.2017.01.014

51. Kresovich JK, Xu Z, O’Brien KM, Weinberg CR, Sandler DP, Taylor JA. Epigenetic mortality predictors and incidence of breast cancer. Aging. Dec 17 2019;11(24):11975–11987. doi:10.18632/aging.102523

52. Chen L, Ganz PA, Sehl ME. DNA Methylation, Aging, and Cancer Risk: A Mini-Review. Front Bioinform. 2022;2:847629. doi:10.3389/fbinf.2022.847629

53. Mullins MA, Kler JS, Eastman MR, Kabeto M, Wallner LP, Kobayashi LC. Validation of Selfreported Cancer Diagnoses Using Medicare Diagnostic Claims in the US Health and Retirement Study, 2000-2016. Cancer Epidemiol Biomarkers Prev. Jan 2022;31(1):287–292. doi:10.1158/1055-9965.Epi-21-0835

