## Supplemental Table 1 for "Aging measures and cancer: Findings from the Health and Retirement Study"

Supplemental Table 1. Description of aging constructs

| Aging constructs | How was the aging construct created in published paper? | Number of biomarkers | Unit | Reference |
|---|---|---|---|---|
| **SA** | Participants responded to the question "How old do you feel?" | NA | Years | |
| | ***Clinical marker-based aging constructs*** | | | |
| **KDM-BA** | Klemara Doubal algorithm was applied to train 10 fairly standard clinical markers in order to constructs KDM- BA. Those 10 markers, which were significantly correlated with age (r>0.1) in the third National Health and Nutrition Examination Survey (NHANES III), were selected from a group of 21 biomarkers known to play a role in the aging process. | 10 clinical markers | Years | Levine [2013][1] |
| **PhenoAge** | Cox penalized regression was applied to train clinical biomarkers and age against mortality. | 9 clinical markers and age | Years | Levine [2018][2] |
| | ***Epigenetc clocks (ECs)*** | | | |
| **Hannum EC** | Elastic net regression combined with bootstrap approaches was applied to train DNA methylation (DNAm) sites, gender, and body mass index (BMI) against age. First generation EC. | 71 CpGs | Years | Hannum [2013][3] |
| **Horvath EC** | Elastic net regression was applied to train DNAm sites against age. First generation EC. | 353 CpGs | Years | Horvath [2013][4] |
| **Levine EC** | Elastic net regression was applied to train DNAm sites against PhenoAge. Second generation EC. | 513 CpGs | Years | Levine [2018][2] |
| **GrimAge** | Elastic net Cox regression was applied to train a DNAm-based composite constructs for seven plasma proteins, smoking pack-years, age and gender against mortality. Second generation EC. | 1030 CpGs | Years | Lu [2019][5] |
| **POA** | Elastic net regression was applied to train DNA methylation sites against the change in 18-biomarker Pace of Aging. Third generation EC. | 46 CpGs | Years of physiological decline per one chronological age | Belsky [2020][6] |


References:

1. Levine ME. Modeling the rate of senescence: can estimated biological age predict mortality more accurately than chronological age? *J Gerontol A Biol Sci Med Sci*. Jun 2013;68(6):667-74. doi:10.1093/gerona/gls233

2. Levine ME, Lu AT, Quach A, et al. An epigenetic biomarker of aging for lifespan and healthspan. *Aging (Albany NY)*. 04 2018;10(4):573-591. doi:10.18632/aging.101414

3. Hannum G, Guinney J, Zhao L, et al. Genome-wide methylation profiles reveal quantitative views of human aging rates. *Mol Cell*. Jan 2013;49(2):359-367. doi:10.1016/j.molcel.2012.10.016

4. Horvath S. DNA methylation age of human tissues and cell types. *Genome Biol*. 2013;14(10):R115. doi:10.1186/gb-2013-14-10-r115

5. Lu AT, Quach A, Wilson JG, et al. DNA methylation GrimAge strongly predicts lifespan and healthspan. *Aging (Albany NY)*. 01 2019;11(2):303-327. doi:10.18632/aging.101684

6. Belsky DW, Caspi A, Arseneault L, et al. Quantification of the pace of biological aging in humans through a blood test, the DunedinPoAm DNA methylation algorithm. *Elife*. 05 2020;9doi:10.7554/eLife.54870
