## Supplemental Table 2 for "Aging measures and cancer: Findings from the Health and Retirement Study"

Supplemental Table 2a. Correlations across KDM-BA, PhenoAge, SA, and chronological age (CA) in Sample A[a]

| | CA | KDM-BA | PhenoAge | SA |
|---|---|---|---|---|
| CA | 1 | | | |
| KDM-BA | 0.92 | 1 | | |
| PhenoAge | 0.80 | 0.84 | 1 | |
| SA | 0.60 | 0.56 | 0.53 | 1 |

[a]Sample A includes 5,501 participants who had reported their SA and had biomarker measures used to calculate KDM-BA and PhenoAge.

Supplemental Table 2b. Correlations between age acceleration (abbreviated as Accel) for KDM-BA, PhenoAge, and SA and chronological age (CA) in Sample A[a]

| | CA | KDM-BA-Accel | PhenoAgeAccel | SA-Accel |
|---|---|---|---|---|
| CA | 1 | | | |
| KDM-BA-Accel | 0 | 1 | | |
| PhenoAgeAccel | 0 | 0.42 | 1 | |
| SA-Accel | 0 | 0.04 | 0.09 | 1 |

[a]Sample A includes 5,501 participants who had reported their SA and had biomarker measures used to calculate KDM-BA and PhenoAge.
