## Supplemental Table 3 for "Aging measures and cancer: Findings from the Health and Retirement Study"

Supplemental Table 3a. Correlations between chronological age (CA) and ECs in Sample B[a]

| Aging constructs | CA | Hannum EC | Horvath EC | Levine EC | GrimAge | POA |
|---|---|---|---|---|---|---|
| CA | 1 | | | | | |
| Hannum EC | 0.81 | 1 | | | | |
| Horvath EC | 0.73 | 0.77 | 1 | | | |
| Levine EC | 0.73 | 0.75 | 0.65 | 1 | | |
| GrimAge | 0.83 | 0.76 | 0.65 | 0.74 | 1 | |
| POA | 0.03 | 0.12 | 0.10 | 0.20 | 0.38 | 1 |

[a]Sample B includes 3,387 participants who had data on ECs.

Supplemental Table 3b. Correlations between CA and age acceleration (abbreviated as Accel) for ECs in Sample B[a]

| Aging constructs | CA | HannumAccel | HorvathAccel | LevineAccel | GrimAgeAccel | POA[b] |
|---|---|---|---|---|---|---|
| CA | 1 | | | | | |
| HannumAccel | 0 | 1 | | | | |
| HorvathAccel | 0 | 0.43 | 1 | | | |
| LevineAccel | 0 | 0.40 | 0.26 | 1 | | |
| GrimAgeAccel | 0 | 0.25 | 0.10 | 0.34 | 1 | |
| POA | 0.03 | 0.16 | 0.12 | 0.25 | 0.63 | 1 |

[a]Sample B includes 3,387 pariticpnats who had data on ECs.
[b]We did not calculate age acceleration for POA because it was not correlated with CA.
