## Supplemental Table 4 for "Aging measures and cancer: Findings from the Health and Retirement Study"

Supplemental Table 4. Associations between aging constructs and cancer prevalence in 2016; cancer cases diagnosed within two years of blood collection were excluded, HRS

| **Sample A**[a] | | | | |
|---|---|---|---|---|
| Aging constructs | No. of cancer survivors[b] | No. of controls[b] | OR (95% CI) per 1 SD increase in aging constructs | |
| | | | Model 1[c] | Model 2[d] |
| KDM-BA-Accel (SD = 3.81 years) | 845 | 4,461 | 1.05 (0.95, 1.15) | 1.00 (0.90, 1.11) |
| PhenoAgeAccel (SD = 7.11 years) | | | 1.14 (1.04, 1.25) | 1.08 (0.97, 1.19) |
| SA-Accel (SD = 10.37 years) | | | 1.17 (1.08, 1.28) | 1.14 (1.04, 1.24) |
| **Sample B**[a] | | | | |
| Aging constructs | No. of cancer survivors[b] | No. of controls[b] | OR (95% CI) per 1 SD increase in aging constructs | |
| | | | Model 1[c] | Model 2[d] |
| HannumAccel (SD = 5.20 years) | 513 | 2,733 | 1.24 (1.10, 1.39) | 1.22 (1.09, 1.37) |
| HorvathAccel (SD = 6.38 years) | | | 1.14 (0.99, 1.30) | 1.12 (0.98, 1.28) |
| LevineAccel (SD = 6.74 years) | | | 1.15 (1.03, 1.28) | 1.12 (1.00, 1.26) |
| GrimAgeAccel (SD = 4.65 years) | | | 1.23 (1.10, 1.39) | 1.21 (1.05, 1.41) |
| POA (SD = 0.09 years of physiological decline per one chronological year) | | | 1.14 (0.99, 1.31) | 1.11 (0.95, 1.29) |

[a]Sample A includes participants who had measures of KDM-BA, PhenoAge, and SA. Sample B includes participants who data on ECs.
[b]Controls are participants without a history of cancer in 2016. Cancer survivors are participants with a history of cancer in 2016.
[c]Model 1 was adjusted for chronological age.
[d]Model 2 was adjusted for chronological age, sex, race/ethnicity, BMI, smoking status, comorbidity index, and CMV infection.
