## Supplemental Table 5 for "Aging measures and cancer: Findings from the Health and Retirement Study"

Supplemental Table 5. Associations between aging constructs and mortality in cancer survivors and controls; cancer cases diagnosed within two years of blood collection were excluded, HRS (2016-2020)

| Aging constructs | No. of deaths | Total person-year | HR (95% CI) per 1 SD increase in aging constructs | |
|---|---|---|---|---|
| **Sample A[a]** | | | | |
| *Cancer survivors (N = 845)[b]* | | | | |
| Aging constructs | No. of deaths | Total person-year | Model 2[d] | Model 2[d] |
| KDM-BA-Accel (SD = 4.02 years) | 108 | 3,379 | 1.14 (0.95, 1.36) | 1.08 (0.89, 1.30) |
| PhenoAgeAccel (SD = 7.19 years) | | | 1.47 (1.13, 1.91) | 1.44 (1.07, 1.94) |
| SA-Accel (SD = 10.33 years) | | | 0.99 (0.68, 1.43) | 0.93 (0.65, 1.34) |
| *Controls (N = 4,461)[b]* | | | | |
| Aging constructs | No. of deaths | Total person-year | Model 2[d] | Model 2[d] |
| KDM-BA-Accel (SD = 3.77 years) | 242 | 18,468 | 1.39 (1.20, 1.61) | 1.32 (1.10, 1.59) |
| PhenoAgeAccel (SD = 7.09 years) | | | 1.46 (1.29, 1.66) | 1.41 (1.21, 1.65) |
| SA-Accel (SD = 10.37 years) | | | 1.24 (0.98, 1.55) | 1.20 (0.98, 1.46) |
| **Sample B[a]** | | | | |
| *Cancer survivors (N = 513)[b]* | | | | |
| Aging constructs | No. of deaths | Total person-year | HR (95% CI) per 1 SD increase in Aging constructs: Model 2[d] | Model 2[d] |
| HannumAccel (SD = 5.48 years) | 88 | 2,011 | 1.23 (1.04, 1.45) | 1.34 (1.13, 1.58) |
| HorvathAccel (SD = 6.94 years) | | | 1.19 (0.95, 1.49) | 1.27 (1.02, 1.58) |
| LevineAccel (DNAmPhenoAge) (SD = 6.89 years) | | | 1.48 (1.22, 1.80) | 1.55 (1.26, 1.93) |
| GrimAgeAccel (SD = 4.66 years) | | | 2.01 (1.55, 2.60) | 2.09 (1.45, 3.00) |
| POA (SD = 0.09 years of physiological decline per one chronological year) | | | 1.30 (1.03, 1.65) | 1.13 (0.82, 1.57) |
| *Controls (N = 2,733)[b]* | | | | |
| Aging constructs | No. of deaths | Total person-year | HR (95% CI) per 1 SD increase in Aging constructs: Model 1[c] | Model 2[d] |
| HannumAccel (SD = 5.11 years) | 207 | 11,145 | 1.22 (1.03, 1.44) | 1.20 (1.00, 1.44) |
| HorvathAccel (SD = 6.23 years) | | | 1.08 (0.88, 1.32) | 1.05 (0.85, 1.31) |
| LevineAccel (SD = 6.72 years) | | | 1.15 (0.94, 1.39) | 1.10 (0.89, 1.34) |
| GrimAgeAccel (SD = 4.65 years) | | | 1.54 (1.32, 1.79) | 1.42 (1.15, 1.74) |
| POA (SD = 0.09 years of physiological decline per one chronological year) | | | 1.42 (1.26, 1.62) | 1.30 (1.14, 1.49) |

[a]Sample A includes participants who had measures of KDM-BA, PhenoAge, and SA. Sample B includes participants who had data on ECs.
[b]Controls are participants without a history of cancer in 2016. Cancer survivors are participants with a history of cancer in 2016.
[c]Model 1 was adjusted for chronological age.
[d]Model 2 was adjusted for chronological age, sex, race/ethnicity, BMI, smoking status, comorbidity index, and CMV infection.
