## Supplemental Table 6 for "Aging measures and cancer: Findings from the Health and Retirement Study"

Supplemental Table 6. Associations between aging constructs and mortality in cancer survivors stratified by sex, HRS (2016-2020)

| **Sample A[a]: 946 cancer survivors** | | | |
|---|---|---|---|
| | **HR (95% CI)[b] per 1 SD increase in age acceleration** | | |
| Sex group | KDM-BA-Accel | PhenoAgeAccel | SA-Accel |
| Female (59 deaths out of 511 survivors) | 1.26 (0.96, 1.65) | 1.50 (1.12, 2.01) | 0.82 (0.52, 1.31) |
| Male (63 deaths out of 435 survivors) | 1.06 (0.77, 1.50) | 1.54 (0.99, 2.40) | 0.92 (0.62, 1.38) |
| P-interaction | 0.44 | 0.87 | 0.60 |
| **Sample B[a]: 582 cancer survivors** | | | |
| | **HR (95% CI)[b] per 1 SD increase in age acceleration** | | |
| Sex group | HannumAccel | HorvathAccel | LevineAccel |
| Female (48 deaths out of 314 survivors) | 1.51 (1.26, 1.81) | 1.37 (1.01, 1.87) | 1.65 (1.35, 2.01) |
| Male (55 deaths out of 268 survivors) | 1.12 (0.86, 1.45) | 1.09 (0.77, 1.53) | 1.55 (1.18, 2.03) |
| P-interaction | 0.27 | 0.55 | 0.99 |
| Sex group | GrimAgeAccel | POA | |
| Female (48 deaths out of 314 survivors) | 2.97 (2.21, 4.00) | 1.44 (1.00, 2.07) | |
| Male (55 deaths out of 268 survivors) | 1.55 (1.03, 2.33) | 1.09 (0.77, 1.54) | |
| P-interaction | 0.0017 | 0.06 | |

[a]Sample A includes cancer survivors who had measures of KDM-BA, PhenoAge, and SA. Sample B includes cancer survivors who had data on ECs.
[b]The model was adjusted for chronological age, race/ethnicity, BMI, smoking status, comorbidity index, and CMV infection.
