## Supplemental Tabel 7 for "Aging measures and cancer: Findings from the Health and Retirement Study"

Supplemental Table 7. Associations between aging constructs and mortality in cancer survivors stratified by time since cancer diagnosis, HRS (2016-2020)

| **Sample A: 941 cancer survivors (5 survivors were excluded due to missing time of cancer diagnosis)** | | | |
|---|---|---|---|
| | **HR (95% CI)[b] per 1 SD increase in age acceleration** | | |
| Time since cancer diagnosis | KDM-BA-Accel | PhenoAgeAccel | SA-Accel |
| <2 years (14 deaths out of 95 survivors) | 1.35 (0.80, 2.31) | 1.63 (0.79, 3.37) | 1.07 (0.56, 2.09) |
| 2-5 years (25 deaths out of 162 survivors) | 1.02 (0.70, 1.46) | 1.90 (1.07, 3.38) | 0.91 (0.49, 1.71) |
| >5 years (83 deaths out of 683 survivors) | 1.09 (0.88, 1.35) | 1.34 (0.96, 1.86) | 0.89 (0.62, 1.27) |
| **Sample B: 577 cancer survivors (5 survivors were excluded due to missing time of cancer diagnosis)** | | | |
| | **HR (95% CI)[b] per 1 SD increase in age acceleration** | | |
| Time since cancer diagnosis | HannumAccel | HorvathAccel | LevineAccel |
| <2 years (15 deaths out of 64 survivors) | 1.33 (0.94, 1.86) | 1.17 (0.88, 1.54) | 3.84 (1.28, 11.54) |
| 2-5 years (15 deaths out of 110 survivors) | 1.32 (0.81, 2.61) | 1.52 (0.98, 2.36) | 2.41 (0.97, 6.01) |
| >5 years (73 deaths out of 403 survivors) | 1.48 (1.14, 1.93) | 1.23 (0.93, 1.61) | 1.49 (1.21, 1.82) |
| Time since cancer diagnosis | GrimAgeAccel | POA | |
| <2 years (15 deaths out of 64 survivors) | 2.77 (1.98, 3.89) | 1.93 (1.49, 2.51) | |
| 2-5 years (15 deaths out of 110 survivors) | 3.08 (1.34, 7.07) | 1.72 (0.35, 8.37) | |
| >5 years (73 deaths out of 403 survivors) | 2.04 (1.30, 3.18) | 1.10 (0.77, 1.58) | |

[a]Sample A includes cancer survivors who had measures of KDM-BA, PhenoAge, and SA. Sample B includes cancer survivors who had data on ECs.
[b]The model was adjusted for chronological age, sex, race/ethnicity, BMI, smoking status, comorbidity index, and CMV infection.
