## Supplemental Table 8 for "Aging measures and cancer: Findings from the Health and Retirement Study"

Supplemental Table 8. Association between aging constructs and risk of total cancer, HRS (2016-2020)

| Aging constructs | No. of cancer cases | Total person-years | HR (95% CI) per 1 SD increase in aging constructs | |
|---|---|---|---|---|
| **Sample A: 4,413 participants without a history of cancer in 2016[a]** | | | | |
| | | | Model 1[b] | Model 2[c] |
| KDM-BA-Accel (SD = 3.76 years) | 182 | 17,405 | 1.05 (0.89, 1.23) | 1.00 (0.84, 1.20) |
| PhenoAgeAccel (SD = 7.07 years) | | | 1.06 (0.88, 1.28) | 0.95 (0.76, 1.18) |
| SA-Accel (SD = 10.35 years) | | | 1.00 (0.82, 1.22) | 0.98 (0.82, 1.16) |
| **Sample B: 2,688 respondents without a history of cancer in 2016[a]** | | | | |
| Aging constructs | No. of cancer cases | Total person-years | HR (95% CI) per 1 SD increase in aging constructs | |
| | | | Model 1[b] | Model 2[c] |
| HannumAccel (SD = 5.13 years) | 122 | 10,366 | 1.15 (0.95, 1.40) | 1.12 (0.90, 1.40) |
| HorvathAccel (SD = 6.26 years) | | | 1.06 (0.88, 1.29) | 1.04 (0.87, 1.26) |
| LevineAccel (SD = 6.77 years) | | | 1.07 (0.86, 1.33) | 1.05 (0.83, 1.32) |
| GrimAgeAccel (SD = 4.64 years) | | | 1.19 (0.98, 1.45) | 1.09 (0.81, 1.45) |
| POA (SD = 0.09 years of physiological decline per one chronological year) | | | 1.23 (0.99, 1.03) | 1.14 (0.81, 1.59) |

[a]Sample A includes 4,413 participants who had measures of KDM-BA, PhenoAge, and SA. Sample B includes 2,688 participants who had data on ECs. Participants with a prevalent cancer in 2016 and with missing cancer status after 2016 or missing cancer diagnosis time were excluded.

[b]Model 1 was adjusted for chronological age.

[c]Model 2 was adjusted for chronological age, sex, race/ethnicity, BMI, smoking status, comorbidity index, and CMV infection.
