## Supplemental Table 9 for "Aging measures and cancer: Findings from the Health and Retirement Study"

Supplemental Table 9a. Correlations between aging constructs and chronological age (CA) in the overlap of participants who were included in both Sample A and Sample B[a] (N = 2,413)

| | CA | KDM-BA | PhenoAge | SA | Hannum EC | Horvath EC | Levine EC | GrimAge | POA |
|---|---|---|---|---|---|---|---|---|---|
| CA | 1 | | | | | | | | |
| KDM-BA | 0.91 | 1 | | | | | | | |
| PhenoAge | 0.80 | 0.84 | 1 | | | | | | |
| SA | 0.61 | 0.57 | 0.53 | 1 | | | | | |
| Hannum EC | 0.81 | 0.76 | 0.70 | 0.52 | 1 | | | | |
| Horvath EC | 0.72 | 0.66 | 0.61 | 0.44 | 0.76 | 1 | | | |
| Levine's EC | 0.72 | 0.68 | 0.66 | 0.45 | 0.75 | 0.64 | 1 | | |
| GrimAge | 0.82 | 0.79 | 0.77 | 0.52 | 0.76 | 0.63 | 0.72 | 1 | |
| POA | 0.05 | 0.09 | 0.22 | 0.06 | 0.14 | 0.12 | 0.21 | 0.40 | 1 |

[a]Participants who were in both Sample A and Sample B had data on all aging constructs.

Supplemental Table 9b. Correlations between age acceleration (abbreviated as Accel) for aging constructs and chronological age (CA) in participants who were included in both Sample A and Sample B[a] (N = 2,413)

| | CA | KDM-BA-Accel | PhenoAgeAccel | SA-Accel | HannumAccel | HorvathAccel | LevineAccel | GrimAgeAccel | POA |
|---|---|---|---|---|---|---|---|---|---|
| CA | 1 | | | | | | | | |
| KDM-BA-Accel | 0.002 | 1 | | | | | | | |
| PhenoAgeAccel | 0.007 | 0.43 | 1 | | | | | | |
| SA-Accel | 0.017 | 0.03 | 0.09 | 1 | | | | | |
| HannumAccel | 0.018 | 0.06 | 0.16 | 0.04 | 1 | | | | |
| HorvathAccel | 0.001 | 0.02 | 0.08 | 0.004 | 0.43 | 1 | | | |
| LevineAccel | 0.003 | 0.09 | 0.20 | 0.03 | 0.42 | 0.26 | 1 | | |
| GrimAgeAccel | 0.002 | 0.15 | 0.33 | 0.03 | 0.26 | 0.11 | 0.33 | 1 | |
| POA | 0.05 | 0.11 | 0.29 | 0.04 | 0.17 | 0.12 | 0.25 | 0.63 | 1 |

[a]Participants who were in both Sample A and Sample B had data on all aging constructs.
