## Supplemental Table 10 for "Aging measures and cancer: Findings from the Health and Retirement Study"

Supplemental Table 10. Association between aging constructs and mortality among cancer survivors[a] and controls[a]; putting eight aging constructs in a single model, HRS (2016-2020)

| *Cancer survivors (N = 410)* | |
|---|---|
| Aging constructs | HR (95% CI)[b] per 1 SD for aging constructs |
| KDM-BA-Accel | 1.24 (0.83, 1.86) |
| PhenoAgeAccel | 1.34 (0.97, 1.85) |
| SA-Accel | 0.88 (0.51, 1.52) |
| HannumAccel | 1.15 (0.86, 1.47) |
| HorvathAccel | 1.16 (0.86, 1.56) |
| LevineAccel | 1.41 (1.07, 1.87) |
| GrimAgeAccel | 1.96 (1.09, 3.53) |
| POA | 0.77 (0.50, 1.21) |
| *Controls (N = 2003)* | |
| Aging constructs | HR (95% CI)[b] per 1 SD for aging constructs |
| KDM-BA-Accel | 1.07 (1.02, 1.13) |
| PhenoAgeAccel | 1.01 (0.98, 1.04) |
| SA-Accel | 1.01 (0.97, 1.04) |
| HannumAccel | 1.01 (0.95, 1.07) |
| HorvathAccel | 1.01 (0.95, 1.07) |
| LevineAccel | 1.04 (0.99, 1.09) |
| GrimAgeAccel | 1.02 (0.95, 1.08) |
| POA | 6.49 (0.81, 52.53) |

[a]Cancer survivors and controls who were in both Sample A and Sample B had data on all aging constructs.

[b]The model was adjusted for chronological age, race/ethnicity, BMI, smoking status, comorbidity index, and CMV infection.
